# Evaluating the Effectiveness of the Mozambique-Canada Maternal Health (MCMH) Project Abstraction Tool in the Identification of Maternal Near-Miss (MNM) Events

**DOI:** 10.1101/2024.05.14.24307349

**Authors:** Maud Z Muosieyiri, Fernanda Andre, Jessie Forsyth, Ana Paula Ferrão da Silva Adoni, Nazeem Muhajarine

**Affiliations:** Department of Community Health and Epidemiology, College of Medicine, University of Saskatchewan, Saskatoon, Saskatchewan, Canada; Department of Biostatistics and Epidemiology, University of Massachusetts, Amherst (UMASS Amherst), Amherst, MA, United States of America; Mozambique-Canada Maternal Health Project, Inhambane Province, Mozambique; Provincial Hospital of Inhambane, Mozambique

## Abstract

Maternal Near-Miss (MNM) is described as a woman who survives a severe obstetric event. The World Health Organization (WHO) developed an abstraction tool in 2009 for identifying MNMs, but it has come under criticism for not being suitable for use in low-resource settings. The maternal near-miss tool developed by the Mozambique-Canada Maternal Health Project, including additional clinical criteria, is an adaptation of the WHO version to suit the resource availability in Mozambique. This study examined whether these additional criteria enhanced maternal near-miss identification; if so, whether this was observed in particular groups of women.

A cross-sectional study was conducted in two hospitals, the Provincial Hospital of Inhambane province, a tertiary referral care center, and a rural hospital, Vilankulo Rural Hospital, with a large rural catchment area (approximately 46,543 inhabitants), in the Inhambane province in Mozambique. Consecutive admissions in the maternity wards in these two hospitals between August 2021 and February 2022 were eligible and data from 2057 women were included. Chi-square test of independence, kappa statistics, and multiple logistic regression analyses were performed to address the study aims.

The newer tool with additional clinical criteria identified more maternal near-misses (Expanded Disease criterion = 28.2%; Comorbidities criterion = 21.1%) than the original WHO tool (16.20%). Hypertension and Anemia, two criteria in the newer tool, showed strong associations with the original WHO disease criterion (p < 0.001). Hypertension demonstrated a moderate agreement with the WHO disease criterion (κ = 0.58, 95% CI: 0.53-0.63) while anemia showed a fair agreement (κ = 0.21, 95% CI: 0.16-0.26). However, HIV/AIDS, the most prevalent comorbidity, was not significantly associated with the original WHO disease criterion. Furthermore, socio-demographic indicators like distance from home to hospital, age of woman, and type of health facility (provincial or rural district) were significant predictors of identifying maternal near-misses.

In conclusion, incorporating additional criteria enhances – it casts a larger net – the original WHO disease criterion’s capacity to identify maternal near-misses. Distance from home to the hospital and age emerge as strong predictors for recognizing MNMs in Inhambane province.

## Introduction

Maternal mortality remains a pressing global health issue. The United Nations (UN) included maternal health promotion in the Millenium Development Goals (MDGs), to reduce global maternal deaths by 75% between 1990 and 2000, as well as in the Sustainable Development Goals (SDGs), to decrease global maternal deaths to less than 70 per 100,000 births by 2030.[1] Yet, maternal mortality still occurs in about 151 per 100,000 births predominantly within Africa, Asia, and Latin America where a combination of clinical and socio-cultural factors significantly increase the risk of maternal deaths.[2–3]

Although maternal mortality is high, it occurs less frequently than maternal near-misses (MNMs).[4] Maternal near-miss refers to an event where a woman nearly died but survived a complication during pregnancy, childbirth, or within 42 days of termination of pregnancy.[5] MNMs are seen as efficient alternatives, to otherwise maternal mortality, for evaluating and improving the quality of maternal care.[6] An important development that has spurred MNM research and action is the creation of an abstraction tool to help clinicians and researchers identify MNMs.[7]

The WHO’s MNM abstraction tool comprises three main criteria: Disease-based, Intervention-based, and Organ-dysfunction criteria. The Disease-based criterion includes five “potentially life-threatening” clinical indicators marking the initial stage in the pathological continuum that eventually leads to death.[7] The Intervention category contains four hospital-based parameters aiding in predicting and mitigating death risks. Lastly, the Organ-dysfunction criterion consists of seven “life-threatening” system failure markers representing the final stage of this pathological continuum directly causing maternal death.[7–8], (Figure S1). According to the WHO, the organ dysfunction criterion is the most effective at predicting MNMs since it is sensitive enough to identify severe life-threatening cases and specific enough to exclude “unnecessary” obstetric events to achieve a manageable workload for clinicians.[7] Evidence from earlier validation studies supported this theory.[9–10] Thus, researchers consider the criterion as the most “epidemiologically sound criterion to use in predicting MNM cases”.[5]

However, subsequent studies have demonstrated major challenges in relying solely on Organ-dysfunction in low- and middle-income countries (LMICs) due to a lack of appropriate laboratory diagnostic equipment and skilled clinicians.[7,10] Most studies further show that the Disease-based criterion is a better standard for identifying MNMs in resource-limited regions because pregnant women usually present with a combination of clinical indicators, due to substantial delays in obtaining obstetric care, that significantly increase their risk of MNMs.[3,9,11] Overwhelming evidence shows hemorrhage, anemia, and hypertension are the leading causes of MNMs in low-income settings.[10–13] For these reasons, revisions have been made to the WHO abstraction tool to improve its utility in resource-limited settings.

The adapted sub-Saharan Africa MNM Abstraction (SSA) tool is one of the well-known adaptations of the WHO abstraction tool and forms the foundation for other country-specific versions of the SSA, like the Nigerian abstraction tool.[11] The tool developed by Mozambique-Canada Maternal Health Project (MCMH project) is a further iteration of the Nigerian version (Figure S2). The MCMH abstraction tool maintains all the indicators of both the Intervention and Organ dysfunction criteria; the latter includes cardiac, respiratory, renal, coagulation/hematological, hepatic, neurologic, and uterine dysfunctions. However, it substantially expands the disease-based markers, by introducing the “Expanded Disease” and “Comorbidities” criteria. The Expanded disease criterion elaborates on each marker from the original WHO disease category while the comorbidities criterion includes non-obstetric comorbidities such as HIV/AIDS and malaria. Additionally, it uniquely incorporates specific socio-demographics known to impact maternal morbidity and mortality.[14–25] Thus, the MCMH abstraction tool, constructed by the Mozambique-Canada Maternal Health working group, aims to capture more comprehensive clinical and socio-demographic information. It was utilized in the MNM 1.0 cross-sectional study in two hospitals in the Inhambane province, Mozambique.

Although the adapted sub-Saharan Africa MNM tools (the MCMH abstraction tool is an example) have been used to collect data, no research has been conducted to investigate how the adapted tool enhances MNM identification. This study addressed this gap by analyzing how the MCMH’s additional clinical criteria may improve the capacity of the original WHO disease criterion in identifying MNM cases. It also examined the impact of specific health systems, geographic, and socio-demographic factors like the type of hospital, distance to the health facility, age of woman, education, profession, marital status, and religion, on MNM identification.

## Materials and Methods

### Study Setting, Design, and Population

Eligible participants were women who presented consecutively at two hospitals, Vilankulo Rural Hospital, a rural referral hospital with a large rural catchment area of approximately 46,543 inhabitants, and Inhambane Provincial Hospital, the central tertiary hospital in the Inhambane province, from August 16, 2021, to February 18, 2022, during pregnancy, labor, delivery, or up to 42 days post-partum or termination of pregnancy (including abortion and ectopic pregnancy). A total of 2057 medical records were retrieved, with 1255 participants from Vilankulo Rural Hospital and 802 from Inhambane Provincial Hospital (HPI).

Inhambane province is located on the southern coast of Mozambique with a population of about 1.4 million.[26] Both hospitals are secondary referral facilities, but HPI is a referral center for the entire province, including Vilankulo and other secondary hospitals. Participants were recruited by well-trained health personnel who approached women in the maternity units in both hospitals. Interested women gave consent to access their records and to participate in the study. The trained personnel also collected data through participant interviews and medical records, using the MCMH abstraction tool. Weekly meetings were held to ensure data quality.

### Ethical Consideration

Application for ethical approval for this secondary use of the MNM 1.0 was sought from the University of Saskatchewan Research Ethics Board. Approval was obtained on December 05, 2022 (Approval number: Bio 3774 NER). To ensure the confidentiality of patient information, only de-identified patient information was used. Original approval for the MNM 1.0 study was obtained from the National Committee on Bioethics in Health, Mozambique on May 20, 2019 (Approval number: 234/CNBS/19). Formal written consent was obtained from all participants in the study. For participants younger than 18 years, written assents were obtained from the children while written informed consent was obtained from their parents/guardians.

### Data Collection Tool

The MCMH abstraction tool collected the clinical and socio-demographic information of participants. The information was divided into the following sections respectively: a) hospitalization information b) socio-demographic data c) vitals and pregnancy history d) organ-dysfunction markers e) intervention indicators f) disease-based markers g) expanded disease markers h) comorbidities indicators i) fetal or neonatal outcome in women with delivery (Figure S2).

A maternal near-miss case could fulfill definitions from any or all five criteria (sections [d] through [h]). Thus, fulfillment of each criterion was counted as an MNM identified by that criterion. For example, an HIV-positive woman with severe bleeding and neurological issues admitted to the intensive care unit (ICU) met Disease-based [f], Intervention [e], Organ dysfunction [d], and Comorbidities [h] criteria. Biomedical data in sections [d] through [h] were abstracted exclusively from medical records. Four data collectors and a supervisor, based at each hospital, received training before data collection for each research site. Data quality was ensured through frequent checks for uniformity, completeness, and accuracy. Following data collection, two additional personnel further cleaned the dataset before analysis.

### Data Analysis

Chi-square tests of independence were conducted to examine the relationship between each additional clinical criterion and the original WHO disease criterion. Kappa statistics were utilized to assess the agreement between indicators of the expanded clinical criteria and the original Disease criteria. Agreement levels were interpreted based on predefined thresholds.[27] To explore hospital-specific differences, the data was stratified, and both chi-square tests of independence and kappa statistics were re-run on health facility-specific data.

To determine the association between specific socio-demographic factors and MNM outcome, multivariable logistic regressions were performed. Initially, bivariate analyses were conducted to assess the relationship between each independent variable and the outcome. Significant variables (p < 0.05) were included in multivariate analyses. Interaction terms were tested using the 2-log-likelihood method, and confounding effects were evaluated. Goodness-of-fit was assessed using the Hosmer-Lemeshow estimates, with a chi-square value and p >0.05 indicating a good model fit. Odds ratios, 95% confidence intervals, and p-values were reported for each model. Facility-based models were obtained through data stratification and following the same steps. Only statistically significant models per hospital were reported.

## Results

### Study Population Characteristics

Distributions of the population characteristics are presented in Table 1. Approximately half (49.8%) of the women were 24 years or younger and nearly one-third of all study participants were either married or lived maritally with their partners. Most women (68%) stayed close to the study health facilities, within 8 kilometers. Less than half of the population (39%) completed secondary school education and even less than one-tenth completed a bachelors or graduate degree. The majority of these women were unemployed (86.7%) and practiced Christianity (93.7%) (Table 1).

**Table 1.** Characteristics of Study Participants, Maternal Near-miss Study, Inhambane Province, Mozambique, (n=2057)

| Variable | Frequency (%) |
| --- | --- |
| **Hospital admitted (n=2055)** |  |
| Inhambane Provincial Hospital | 800 (38.9) |
| Vilankulo Rural Hospital | 1255 (61.1) |
| **Distance to Health Facility (n=2054)** |  |
| Within 8km to hospital | 1397 (68.0) |
| More than 8km | 657 (32.0) |
| **Educational Level (n=2051)** |  |
| None | 246 (12.0) |
| Primary School 1 | 255 (12.4) |
| Primary School 2 | 545 (26.6) |
| Secondary School | 633 (30.9) |
| Post Secondary | 285 (13.9) |
| Bachelors | 77 (3.7) |
| Graduate | 10 (0.5) |
| **Profession (n=2053)** |  |
| Unemployed | 1735 (86.7) |
| Unqualified Employment | 161 (8.2) |
| Semi-qualified Employment | 92 (4.6) |
| Professional | 11 (0.5) |
| **Religion (n=2045)** |  |
| Islam | 60 (3.1) |
| Christianity | 1867 (93.7) |
| Traditional | 63 (3.1) |
| Other | 2 (0.1) |
| **Marital Status (n=2052)** |  |
| Single | 518 (25.9) |
| Married | 96 (5.0) |
| Live Maritally | 1374 (68.6) |
| Divorced | 9 (0.4) |
| Widow | 2 (0.1) |
| **Age Categories (n=2057)** |  |
| ≤19 | 437 (21.8) |
| 20-24 | 558 (27.8) |
| 25-29 | 433 (21.6) |
| 30-34 | 354 (17.9) |
| 35-39 | 165 (8.2) |
| ≥40 | 55 (2.7) |

### Maternal Near-Misses Identified by Different Criteria

Table 2 and Figure 1 show the distributions of MNMs identified by each criterion. The Expanded disease (28.2%) and Comorbidities criteria (21.1%) identified the highest MNM cases while the Organ-dysfunction criterion yielded the least (2.7%). Each criterion comprises prominent markers that helped identify MNMs for that category (Table 3). Hypertension contributed to the most cases in the original WHO Disease criterion (66.6%) while Infection represented the least (1.5%). Blood transfusion contributed to the most events under the Intervention criterion (71.3%), no patient underwent interventional radiology. Neurological dysfunction accounted for the most cases under the Organ-dysfunction criterion (58.2%), while liver dysfunction showed the least contribution (1.8%). Hypertension remained the highest contributor to the Expanded Disease criterion (36.6%) while infection remained the least (1.6%). MNMs under the Comorbidities criterion were mainly attributed to HIV/AIDS (42.9%), anemia (6.5%), and malaria (1.9%). Medical conditions like kidney, heart, liver, and lung diseases, as well as cancer, were not identified in any study participants.

**Figure 1.**
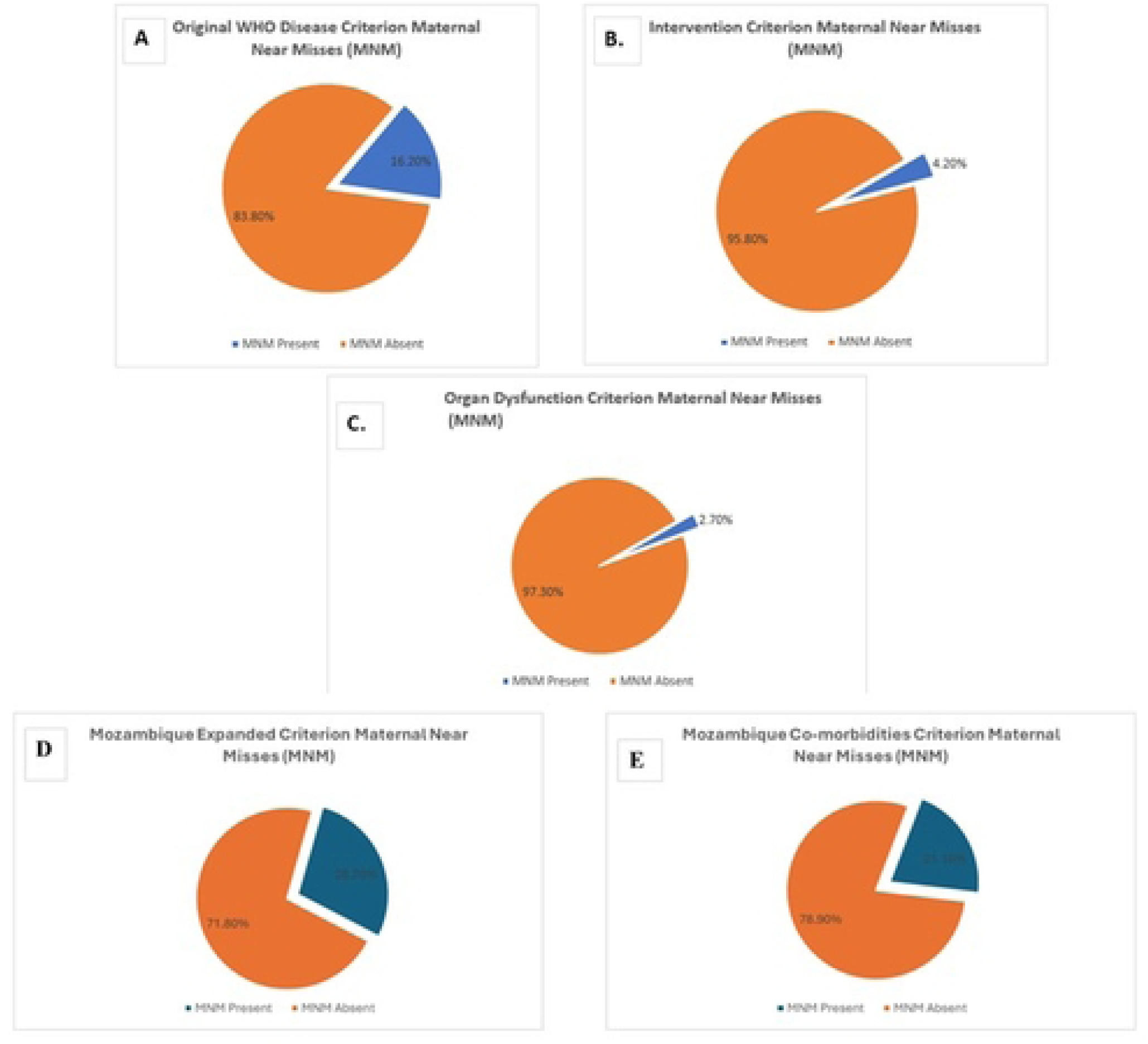
Maternal Near-Misses Identified by the Different Criteria (Fig A – F) in the Mozambique-Canada Maternal Health Abstraction Tool.

**Table 2.** Frequency Distributions of Maternal Near-miss as Identified by Different Criteria, the WHO and Mozambique-Canada Maternal Health Project Abstraction tools (n=2057)

| Criteria | Frequency (%) |
| --- | --- |
| WHO Disease grouping (n = 2054) | 332 (16.2) |
| WHO Interventions grouping (n = 2057) | 87 (4.2) |
| WHO Organ-dysfunction grouping (n = 2055) | 55 (2.7) |
| Mozambique-Canada Maternal Health Tool, Expanded Disease grouping (n = 2057) | 580 (28.2) |
| Mozambique-Canada Maternal Health Tool, Comorbidities grouping (n = 2056) | 434 (21.1) |

**Table 3.**
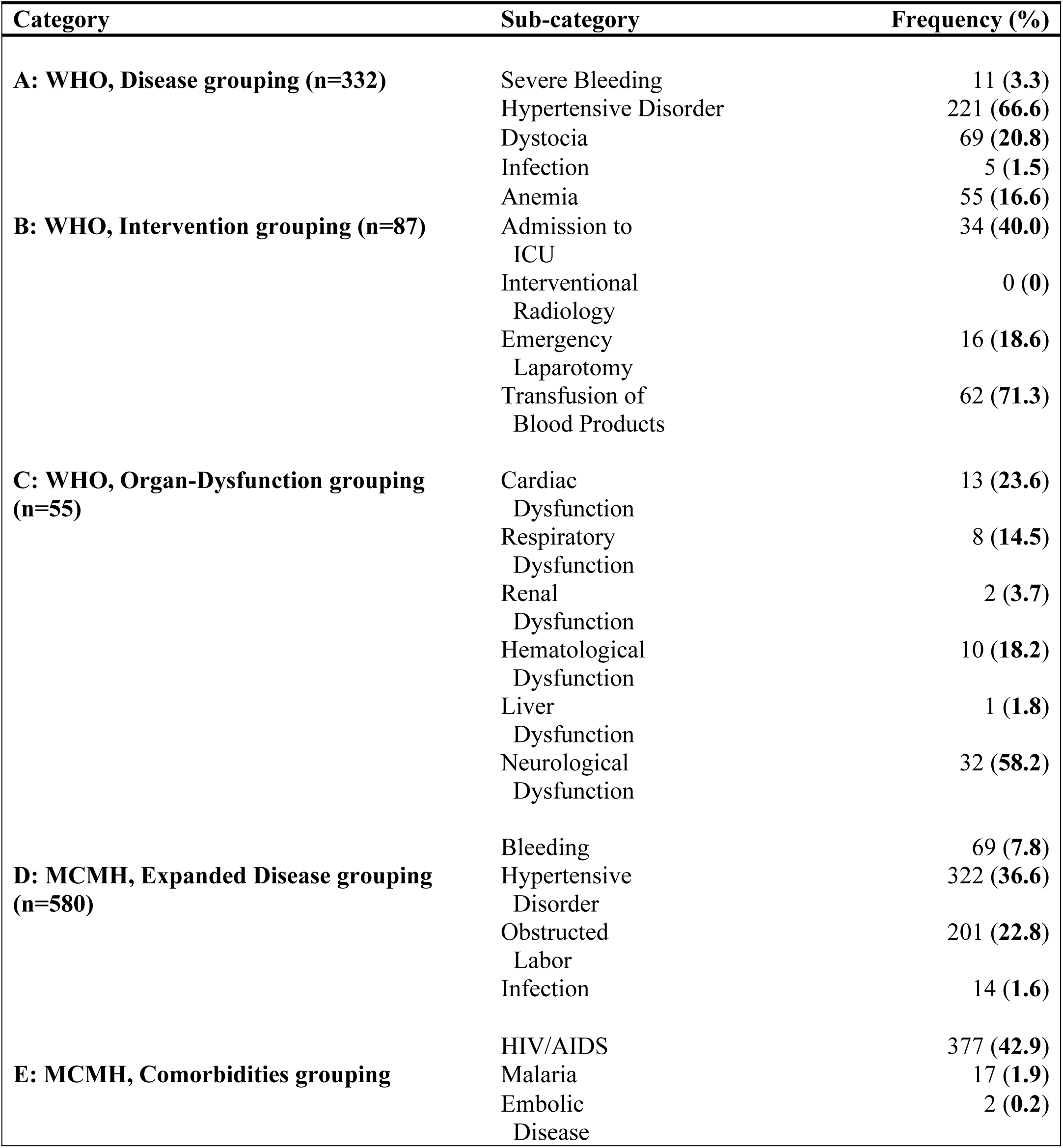
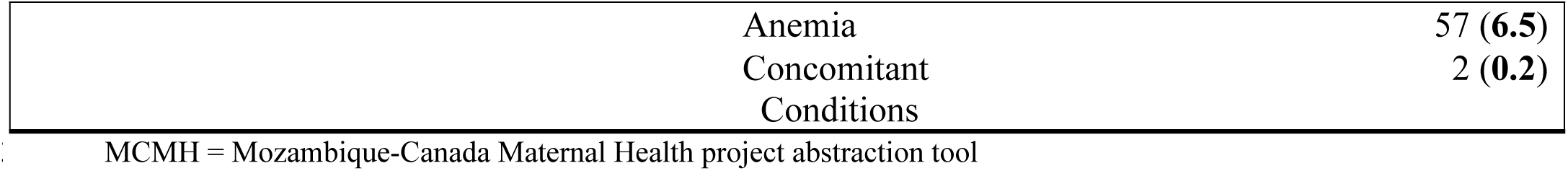
Each Sub-Category Contribution to Maternal Near-miss Identification (n=2057)

| Category | Sub-category | Frequency (%) |
| --- | --- | --- |
| **A: WHO, Disease grouping (n=332)** | Severe Bleeding | 11 (3.3) |
|  | Hypertensive Disorder | 221 (66.6) |
|  | Dystocia | 69 (20.8) |
|  | Infection | 5 (1.5) |
|  | Anemia | 55 (16.6) |
| **B: WHO, Intervention grouping (n=87)** | Admission to ICU | 34 (40.0) |
|  | Interventional Radiology | 0 (0) |
|  | Emergency Laparotomy | 16 (18.6) |
|  | Transfusion of Blood Products | 62 (71.3) |
| **C: WHO, Organ-Dysfunction grouping (n=55)** | Cardiac Dysfunction | 13 (23.6) |
|  | Respiratory Dysfunction | 8 (14.5) |
|  | Renal Dysfunction | 2 (3.7) |
|  | Hematological Dysfunction | 10 (18.2) |
|  | Liver Dysfunction | 1 (1.8) |
|  | Neurological Dysfunction | 32 (58.2) |
| **D: MCMH, Expanded Disease grouping (n=580)** | Bleeding | 69 (7.8) |
|  | Hypertensive Disorder | 322 (36.6) |
|  | Obstructed Labor | 201 (22.8) |
|  | Infection | 14 (1.6) |
| **E: MCMH, Comorbidities grouping** | HIV/AIDS | 377 (42.9) |
|  | Malaria | 17 (1.9) |
|  | Embolic Disease | 2 (0.2) |

|  |  |
| --- | --- |
| Anemia | 57 (6.5) |
| Concomitant Conditions | 2 (0.2) |

MCMH = Mozambique-Canada Maternal Health project abstraction tool

Data from Table 3 was stratified by hospitals (S3 Table). Hypertension, admission to ICU, neurological dysfunction, and HIV/AIDS remained the predominant markers of MNMs in both sites. Notably, MNMs associated with hypertensive disorder (16.2% for WHO Disease, and 20.6% for Expanded Disease) and with ICU admission (2.9%) remained highest among patients in the provincial hospital, while neurological dysfunction (2.1%) and HIV/AIDs (24.2%) were identified as the most prominent cause associated with MNMs in Vilankulo Rural District Hospital.

### Indicators of the Expanded Clinical Criteria that are Associated with the WHO Disease-Criterion

As presented in Table 4, all variables in the Expanded Disease-criterion were statistically associated with the original WHO Disease-criterion (p <0.001). Within this category, hypertension had the strongest association with the original disease category (ꭓ2 = 678.5, [d.f. = 1, p < 0.001]) while obstructed labor had the weakest association with the original WHO Disease criterion (ꭓ2 = 24.0, [d.f. = 1, p < 0.001]). Variables in the Comorbidities category were also statistically related to the original WHO Disease criterion (p < 0.05) with the exception of HIV/AIDS (p > 0.05). Among the statistically significant, anemia was most strongly associated with the original WHO Disease-criterion (ꭓ2 = 200.1, [d.f. = 1, p < 0.001]) while malaria had the weakest association with this original WHO Disease category (ꭓ2 = 4.6, [d.f. = 1, p < 0.005]).

**Table 4.**
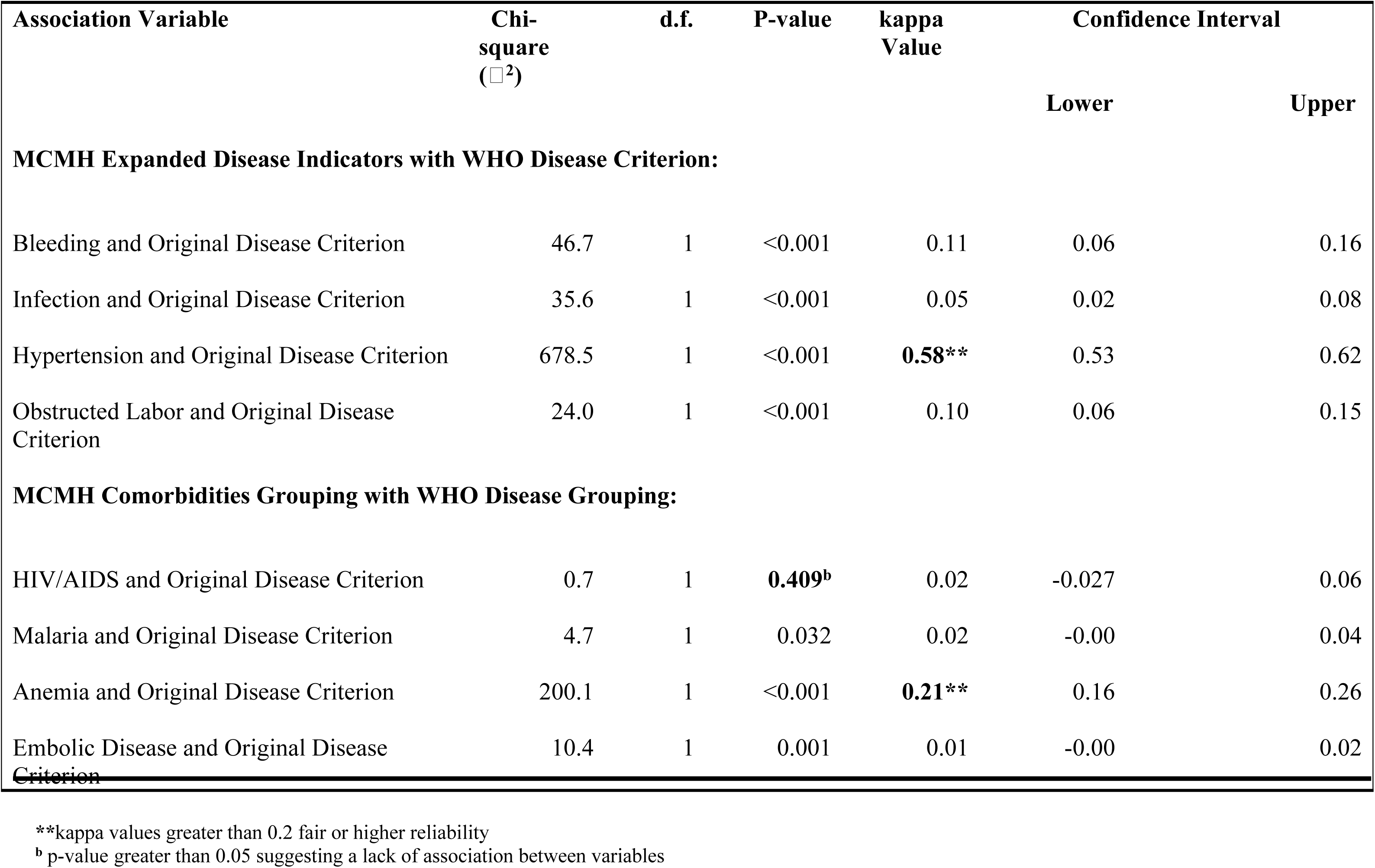
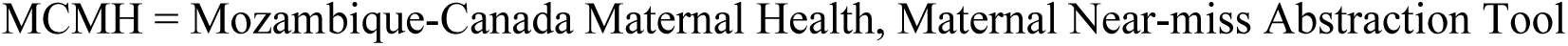
Overall Results from Chi-Square Test of Independence and kappa Statistic Between the WHO Disease Criterion vs the Mozambique-Canada Maternal Health Expanded Disease and Comorbidities Criteria.

| Association Variable | Chi-square<br>($\chi^2$) | d.f. | P-value | kappa<br>Value | Confidence Interval | |
| --- | --- | --- | --- | --- | --- | --- |
|  |  |  |  |  | Lower | Upper |
| MCMH Expanded Disease Indicators with WHO Disease Criterion: |  |  |  |  |  |  |
| Bleeding and Original Disease Criterion | 46.7 | 1 | <0.001 | 0.11 | 0.06 | 0.16 |
| Infection and Original Disease Criterion | 35.6 | 1 | <0.001 | 0.05 | 0.02 | 0.08 |
| Hypertension and Original Disease Criterion | 678.5 | 1 | <0.001 | **0.58\*\*** | 0.53 | 0.62 |
| Obstructed Labor and Original Disease Criterion | 24.0 | 1 | <0.001 | 0.10 | 0.06 | 0.15 |
| MCMH Comorbidities Grouping with WHO Disease Grouping: |  |  |  |  |  |  |
| HIV/AIDS and Original Disease Criterion | 0.7 | 1 | **0.409<sup>b</sup>** | 0.02 | -0.027 | 0.06 |
| Malaria and Original Disease Criterion | 4.7 | 1 | 0.032 | 0.02 | -0.00 | 0.04 |
| Anemia and Original Disease Criterion | 200.1 | 1 | <0.001 | **0.21\*\*** | 0.16 | 0.26 |
| Embolic Disease and Original Disease Criterion | 10.4 | 1 | 0.001 | 0.01 | -0.00 | 0.02 |

\*\*kappa values greater than 0.2 fair or higher reliability
<sup>b</sup> p-value greater than 0.05 suggesting a lack of association between variables
MCMH = Mozambique-Canada Maternal Health, Maternal Near-miss Abstraction Tool

The kappa estimates, which evaluates the corroboration between the original WHO Disease criteria and MCMH Expanded Disease indicators, revealed an overall ‘weak’ agreement between the two categories (Table 4). Only hypertension, one specific condition within MCMH Expanded Disease, showed a ’moderate’ degree of agreement with the original WHO Disease criterion (κ = 0.58, 95% CI: [0.53,0.63]). Generally, the kappa estimates for the MCMH Comorbidities group and WHO Disease criterion were lower than those for the MCMH Expanded Disease criterion and WHO Disease criterion. Most comorbidities showed no agreement with the original WHO Disease criterion, with embolic disease having the lowest level of agreement (κ = 0.01, 95% CI: [0.00, 0.02]). Anemia was the only factor to have a ‘fair’ agreement with the original WHO Disease criterion (κ = 0.21, 95% CI: [0.16, 0.26]).

Similar tests were conducted on hospital-stratified data to identify hospital-specific characteristics. In provincial hospital specific data, hypertension remained strongly associated (ꭓ2 = 336.7, [d.f. = 1, p < 0.001]) and showed an improved agreement, from ’moderate’ to ’substantial’, with the original WHO Disease criterion (κ = 0.65, 95% CI:[0.58, 0.72]. For the comorbidity variables, HIV/AIDS continued to lack an association with the original WHO Disease group (p > 0.05) (S4 Table). In Vilankulo Rural Hospital data, hypertension retained the strongest association with the original WHO Disease category (ꭓ2 = 317.1, [d.f. = 1, p < 0.001]). Hypertension had a ‘moderate’ agreement with the original WHO Disease category (κ = 0.50, 95% CI: [0.43-0.57]) and HIV/AIDS still showed no association with the WHO Disease group (p > 0.05) (S5 Table).

### Socio-demographic Factors that are Associated with Identifying MNMs Defined by Each Clinical Criterion

Table 5 displays results of the multivariable analysis on the relationship between socio- demographic factors and WHO abstraction tool defined MNMs. The first multivariable analysis examined the relationship between socio-demographic factors and the original WHO Disease- criterion defined MNMs. The Hosmer-Lemeshow test indicates a good fit (ꭓ2 = 4.8 [d.f. = 8, p = 0.780 > 0.05]) with the model successfully predicting observed data points (84%). None of the interaction terms tested were statistically significant and no confounding effects were observed. Only distance to the health facility was statistically significant in this model (Figure 2), indicating that individuals living over 8km from the study hospitals had over twice the odds of being identified MNMs compared to those within 8km (OR = 2.47, 95% CI: [1.92,3.18]).

**Figure 2.**
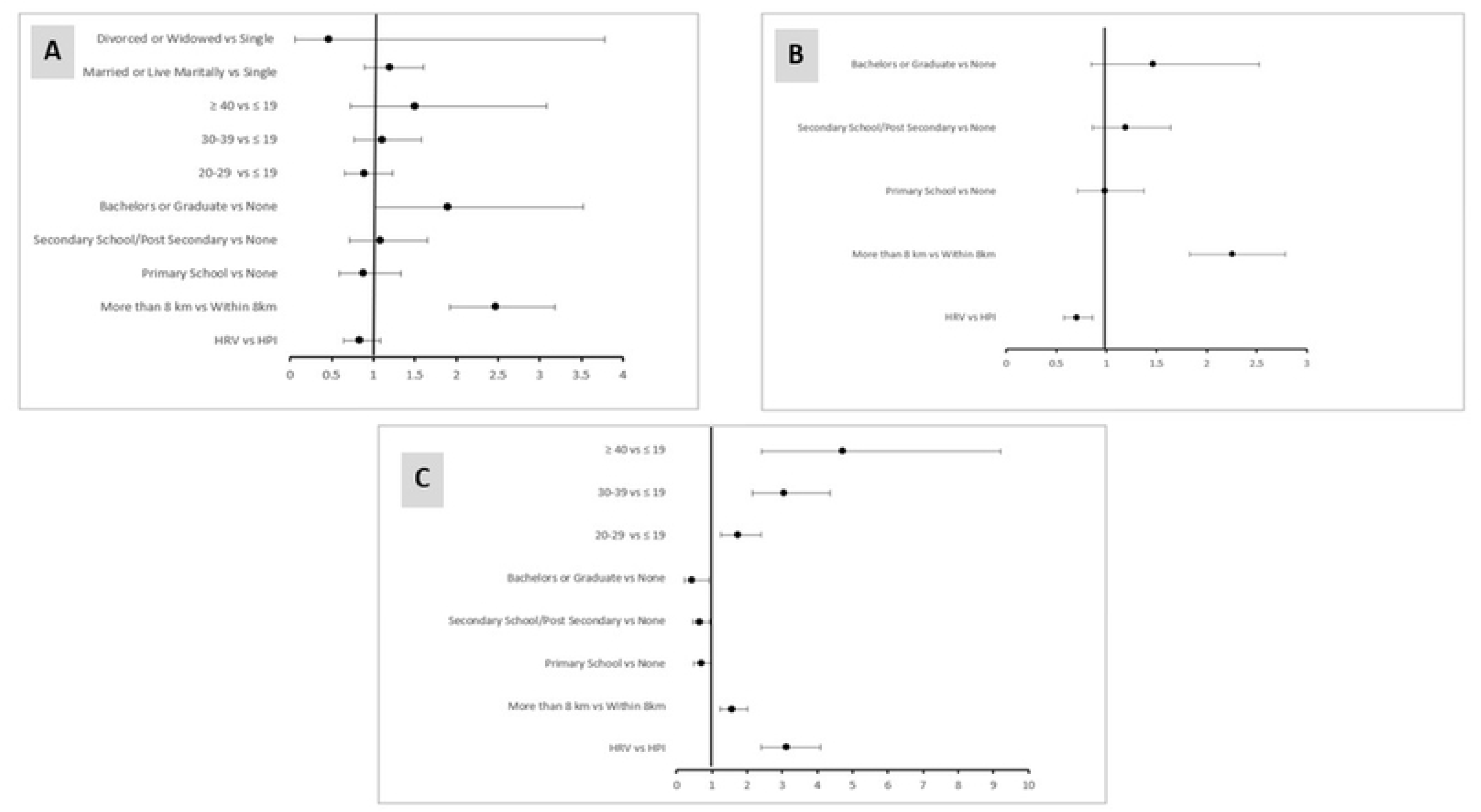
Odds Ratios representing associations between sociodemographic factors and Maternal Near-Misses as defined by the WHO Disease criterion (A), MCMH expanded disease criterion (B), and MCMH co-morbidities criterion (C)

**Table 5.** Multivariable Analysis on the Association Between Maternal Characteristics and WHO Disease Criterion for Identifying MNMs.

| Variables | Adjusted OR | P-Value |
| --- | --- | --- |
| **Hospital admitted** |  |  |
| Inhambane Provincial Hospital | 1 [Ref] |  |
| Vilankulo Rural Hospital | 0.84 [0.65-1.09] | 0.186 |
| **Distance to Health Facility** |  |  |
| Within 8 km | 1 [Ref] |  |
| More than 8 km | 2.47 [1.92-3.18] | <0.001 |
| **Educational Level** |  |  |
| None | 1 [Ref] |  |
| Primary School | 0.88 [0.59-1.33] | 0.554 |
| Secondary School or Post-Secondary | 1.09 [0.72-1.65] | 0.684 |
| Bachelors or Graduate | 1.89 [1.02- 3.52] | **0.043** |
| **Age Categories** |  |  |
| ≤ 19 | 1 [Ref] |  |
| 20-29 | 0.90 [0.66-1.23] | 0.511 |
| 30-39 | 1.11 [0.77-1.59] | 0.589 |
| ≥ 40 | 1.50 [0.73-3.09] | 0.272 |
| **Marital Status** |  |  |
| Single | 1 [Ref] |  |
| Married or Live Maritally | 1.20 [0.89-1.61] | 0.229 |
| Divorced or Widowed | 0.47 [0.06-3.78] | 0.474 |

All boldened p-values indicate statistically significant values (p-value < 0.05) Hosmer-Lemeshow Goodness of fit: ( $\chi^2 = 4.790$ [d.f. = 8, p-value = 0.780 > 0.05])

The second multivariable analysis explored the association between socio-demographic predictors and MCMH Expanded Disease-defined MNMs (Table 6). Hosmer-Lemeshow test showed that the final multivariable model is well-fitted (ꭓ2 = 4.9 [d.f. = 7, p = 0.668 > 0.05]) and has a 71.9% accurate prediction of the observed events. No interaction was determined through statistical testing nor was confounding effects observed. Distance from the health facility remained highly associated with MNM identification, with individuals living over 8km to the hospital having over twice the odds compared to those within 8km (OR = 2.26, 95%CI:1[.83-2.78]). Additionally, the type of hospital was a significant factor, with 29% lower likelihood of identifying an MNM case among the rural hospital cases compared to provincial hospital cases (OR = 0.70, 95%CI: [0.57-0.87]) (Table 6, Figure 2).

**Table 6.** Multivariable Analysis on the Association Between Maternal Characteristics and the Mozambique-Canada Maternal Health Project Expanded Disease Criterion MNMs.

| Variables | Adjusted OR | P-Value |
| --- | --- | --- |
| **Hospital admitted** |  |  |
| Inhambane Provincial Hospital | 1 [Ref] |  |
| Vilankulo District Hospital | 0.70 [0.57-0.87] | **0.001** |
| **Place of Residence** |  |  |
| Within 8 km | 1 [Ref] |  |
| More than 8 km | 2.26 [1.83-2.78] | **<0.001** |
| **Educational Level** |  |  |
| None | 1 [Ref] |  |
| Primary School | 1.00 [0.71-1.37] | 0.946 |
| Secondary School or Post-Secondary | 1.19 [0.87-1.64] | 0.283 |
| Bachelors or Graduate | 1.47 [0.85- 2.52] | 0.167 |

All boldened p-values indicate statistically significant values (p-value < 0.05) Hosmer-Lemeshow Goodness of fit: ( $\chi^2 = 4.937$ [d.f. = 7, p-value = 0.668 > 0.05])

Table 7 shows the results from the third logistic regression model between socio- demographic factors and MCMH Comorbidities-defined MNMs. Results from the Hosmer-Lemeshow test demonstrated that the final multivariable model is well-fitted (ꭓ2 = 5.8 [d.f. = 8, p = 0.674 > 0.05]) with a 79.7% accurate prediction of observed events. No interactions between covariates were observed. However, both maternal educational level and type of hospital produced confounding effects (|Adjusted - Crude ORs| > 10%). In the adjusted model, individuals living over 8km were about 57% more likely to be identified as MNMs than those within that distance (OR = 1.58, 95%CI: [1.24,2.01]). Unlike previous MNM groups (WHO Disease-Criterion defined and MCMH Expanded Disease defined MNMs), admission to the rural hospital, rather than the provincial hospital, increased the odds of identifying comorbidities-defined MNMs (OR = 3.13, 95%CI: [2.40-4.10]) (Table 7, Figure 2).

**Table 7.** Multivariable Analysis on the Association Between Maternal Characteristics and the Mozambique-Canada Maternal Health Project Comorbidities Criterion MNMs.

| Variables | Adjusted OR | P-Value |
| --- | --- | --- |
| **Hospital admitted** |  |  |
| Inhambane Provincial Hospital | 1 [Ref] |  |
| Vilankulo Rural Hospital | 3.13 [2.40-4.08] | **<0.001** |
| **Distance to Health Facility** |  |  |
| Within 8 km | 1 [Ref] |  |
| More than 8 km | 1.58 [1.24-2.01] | **<0.001** |
| **Educational Level** |  |  |
| None | 1 [Ref] |  |
| Primary School | 0.70 [0.49-0.98] | **0.040** |
| Secondary School or Post-Secondary | 0.67 [0.47-0.96] | **0.028** |
| Bachelors or Graduate | 0.45 [0.22- 0.91] | **0.027** |
| **Age Categories** |  |  |
| ≤ 19 | 1 [Ref] |  |
| 20-29 | 1.73 [1.25-2.40] | **<0.001** |
| 30-39 | 3.06 [2.15-4.36] | **<0.001** |
| ≥ 40 | 4.73 [2.43-9.20] | **<0.001** |

All boldened p-values indicate statistical significance of variable (p-value <0.05)
Hosmer-Lemeshow Goodness of fit: ( $\chi^2 = 5.757$ [d.f. = 8, p-value = 0.674 > 0.05])

## Discussion

This study assessed if the additional clinical criteria, Expanded Disease and Comorbidities, as measured by the MNM abstraction tool developed by Mozambique-Canada Maternal Health Project improved the capacity of the original WHO Disease criterion in identifying potential MNMs. It also examined what socio-demographic factors influenced the identification of MNMs in Inhambane, Mozambique.

It is well argued that MNM cases represent the severe end of obstetrical complications; it has been reported that for every maternal death, there may be up to 12 maternal near-miss cases.[29] Therefore, broadly identifying potential MNM cases, especially in low-resource settings, has value clinically. This enables health care providers to be vigilant and to work towards preventing severe obstetrical risks, ideally, or, failing that, to care for those with risk for severe obstetric outcomes and arrest the progression to mortality.

The study findings revealed that the expanded disease criteria identified more MNMs than the original WHO Disease category, with hypertensive disorders being the main contributor to this enhanced identification of MNMs. Both the Expanded Disease and original WHO Disease categories target similar populations, with the former identifying more MNMs. There was less overlap between Comorbidities indicators and the original WHO Disease criterion, except for anemia. The Organ dysfunction criterion was the most conservative for identifying MNMs in the current study. These results are generally consistent with previous literature.[12, 30–31] A clinical implication of these findings is that women who present with chronic hypertensive disorders or anemia are at further risk for severe obstetric outcomes and will need rapid and targeted care to avoid negative outcomes including death.

Similar to the study findings, the sub-Saharan Africa MNM abstraction literature demonstrates that hypertensive disorders is one of the biggest clinical markers of MNMs. Research shows that hypertensive disorders account for about 20% to 53% of all MNM cases in SSA.[32–35] Other studies also include dystocia and anemia as top causes of maternal morbidity.[4,12, 36–38] Although hemorrhage is also reported as a prominent marker for MNMs in SSA, this was not established in our study. Most studies report post-partum hemorrhage as the highest cause of MNMs in SSA ranging from 20% to 57% of all cases.[12–13, 33–35,39–44] However, in the current study, hemorrhage contributed only to 3.3% of all cases, may be partly explained by an effective hospital management protocol for hemorrhage in the hospitals participating in this study, such as the rapid intervention on transfusion of blood or its products.[43] This management protocol is further evidenced by the fact that about 72% of all MNMs identified by the Intervention criterion received a blood transfusion. Alternatively, the low hemorrhage cases could be attributed to a renewed focus on continuous clinical training on managing obstetric hemorrhage, in part, as a result of the training provided by the MCMH project within the Inhambane province.

Although HIV/AIDS was prevalent, it consistently lacked an association with the original WHO Disease category in both hospitals. Other Comorbidity markers, like malaria and embolic disease (thrombo/amniotic/air embolism), showed weak associations with the original WHO Disease group as well. Researchers assert that Comorbid or pre-existing non-obstetric indicators only account for a small subset of all MNMs. For instance, a study showed that only 2.5% of cases were associated with HIV/AIDS and 4.1% with malaria.[44] Oladapo and colleagues [32] noted that these Comorbid diseases contributed only marginally to the overall MNM cases in their study. However, they also observed that these diseases disproportionately contributed more to maternal deaths. Thus, in their study, while only 6.8% of all MNMs were attributed to Comorbid disease, about 19.6% of maternal deaths were associated with these underlying non-obstetric markers.[32] Overall, the results indicate that the Comorbidities criterion, except anemia, does not help improve the capacity of the original WHO Disease criterion in identifying MNMs in both hospitals.

To further understand the profile of a potential MNM, it is necessary to understand the structural factors underpinning the condition. Distance from the hospital was consistently associated with MNMs, regardless of the clinical definition. Thaddeus and Maine stipulated, in their three-delay framework, that distance from a health facility was an essential determinant of the second type of delay that increases the risk of MNM and/or death.[45] In many rural areas within sub-Saharan Africa, the paucity of public transportation, high transport costs, and/or poor road infrastructure exacerbate the delays in reaching these facilities during an obstetric complication.[30,37,46–47] The unavailability of suitable transportation consequently forces some women to walk the distance during an obstetric complication.[46–48] A study showed that walking for more than 1 hour to a health facility was associated with about 4 times higher odds of MNMs.[47] Another study revealed that delays caused by the lack of vehicles increased the odds of MNMs by 8 times.[49] Furthermore, Hadush observed that delays in reaching a health facility contributed to about 40% of the maternal morbidities in their study.[50] Our results reveal similar trends; it suggests that significant delays potentially occur for women who live greater than 8km from a health facility due to transport-related issues which subsequently increases their odds of MNMs.

The type of hospital, essentially a provincial hospital drawing referrals from the entire province or a district hospital with a large rural catchment area, was also significantly associated with the identification of MNMs by utilizing the additional clinical categories. As compared to the provincial hospital, it was less likely for women to be identified as MNMs in the district hospital when using the Expanded Disease criterion. One potential reason is that, as a provincial referral hospital, the Inhambane Provincial Hospital receives women in more critical clinical conditions than those seen at the Vilankulo Rural Hospital. Conversely, the odds of identifying MNMs using the Comorbidities criterion were higher in the rural hospital than in the provincial hospital because the Vilankulo Rural Hospital receives more pregnant women with non-pregnancy related comorbid conditions, such as HIV/AIDS and malaria. It is interesting to note that the type of hospital did not influence the identification of MNMs under the original WHO Disease-criterion. Again, this suggests that the WHO abstraction tool is more conservative in MNM identification.[13] Therefore, incorporating these expanded disease categories possibly expands the range of structural factors to consider, such as the type of hospital, when creating a potential MNM profile.

As expected, older age at delivery increased the odds of identifying MNMs in the Comorbidities criterion.[47,51–54] It is well-established that the risk of maternal morbidities is higher at two periods in a woman’s reproductive life cycle.[26,53] The first is during adolescence (10 years -19 years) and the second is at the end of a woman’s reproductive period (35 years and above.[53,55] One study shows that for women 10 to 15 years, their risk of maternal mortality is about five-fold higher than women between 20 to 24 years.[55] Another study presented that women older than 35 years were 74% more likely to develop a maternal near-miss than women between 25 and 34 years.[51] Women above 35 years especially run a higher risk of developing comorbid disease such as hypertension, heart and thyroid disease, and diabetes that complicate their pregnancies and make them more susceptible to MNMs.[51] This may also explain why the odds of identifying MNMs in women over 35 years was almost 5-fold within the Comorbidities group in our study as well. Although this socio-demographic factor did not show a significant association with MNMs identified by the other clinical criteria, it is important to realize that a similar trend was seen with the odds of identifying MNMs defined by the original WHO Disease criterion. In summary, age may be an important factor to consider when building the MNM profile especially when relying on the Comorbidities definition within secondary hospitals across Inhambane.

Our study did not find any consistent associations between maternal education, marital status, profession or religion and MNMs defined by all clinical criteria. This is potentially because of the homogeneity of the study population in our sample. A majority of women had not completed formal schooling, were married, were unemployed, and declared their religion as Christian. Different groups of women with varying demographics should therefore be included in future studies.

This study has several strengths. To the best of our knowledge, it is one of the first studies that attempt to evaluate an adapted MNM evaluation tool, i.e. MCMH abstraction tool, and to refine the WHO clinical definition of MNMs based on local Mozambican clinical contexts. It also sheds more light on specific socio-demographic factors that help influence MNM identification in Inhambane. Moreover, the design of the MCMH abstraction tool – to include both the additional (Expanded Disease group) and original WHO Disease criterion – helps to better compare data and improve internal validity. This sharply contrasts with other studies where the WHO tool is completely separated and is different in design from the adapted versions, potentially causing inconsistencies in the data collection and results comparison process.[32–33,38,51] Again, this study worked directly with local clinicians who provided relevant clinical and cultural information to help contextualize the investigation. Another major strength was our considerable study sample which was comparable to the sample sizes present in the literature.

Nevertheless, some limitations must be acknowledged. For instance, we observed some data quality issues, such as participants receiving the same unique identifiers on separate hospital admissions. However, these errors were corrected during the data-cleaning phase. Also, the rigor of the study could have been further strengthened by introducing external control. Specifically, a clinician could have performed their clinical diagnoses of potential MNMs, independent of any abstraction criterion, as another source of comparison with both the WHO disease and additional clinical criteria. Another limitation is the homogeneity of the study population which potentially narrowed the ability to broadly generalize the findings beyond this circumscribed population. Despite these limitations, our study unveiled important findings concerning MNM identification in rural Mozambique settings. We hope that these findings can be applied to the current maternal health practices to improve obstetric care, especially within health facilities across the province of Inhambane.

To further advance the identification of MNMs using the MCMH model, future research is needed. For example, more investigation is required to determine if the MCMH abstraction tool produces similar results within different levels of care, such as primary and tertiary-level facilities. Although some socio-demographic factors were not statistically associated with MNMs in our study, further research should be done to test these associations within more diverse study populations. Furthermore, other socio-demographic factors based on the available literature that were not originally captured should also be tested. Finally, qualitative studies should be conducted to provide more insight that complement the findings of this present study.

## Conclusion

This study demonstrated that using expanded clinical categories identified more potential MNM cases than using the WHO abstraction tool, especially the disease group as a criterion. Incorporating markers from an Expanded Disease-criterion, to identify MNMs strongly corroborated the identification of MNMs using the original WHO disease group. Conversely, using markers like HIV/AIDS within the Comorbidities criterion to identify MNMs consistently lacked corroboration with the original WHO clinical group, indicating less overlap between their populations. Therefore, relying on non-pregnancy-related comorbidities, i.e., HIV/AIDS, to profile women at greater risk for MNMs should be done cautiously. Geographical and socio-demographic factors such as longer distance to a health facility and older age of women increased the odds of identifying MNMs.

In summary, the study supports using the Expanded Disease-criterion alongside the original WHO disease category to identify a wider range of MNMs. It also emphasizes assessing specific socio-demographic factors, such as distance to the birth facility and age, to guide the identification of these cases.

## Data Availability

The data that support the findings of this study are openly available in [repository name] at https://doi.org/10.5061/dryad.r2280gbm9

## Acknowledgment

The authors would like to thank members of the Mozambique Maternal Near-Miss Working Group (Stélio Tembe, Renata Munguambe, Jaciara Mussá, Assuçena Maíte, Lídia Mondlane, Milton Moçambique, Emilton Dgedge, David Arone, Joaquim Matshine, Fernanda Sumbana, Dorcia Mandlate) for helpful discussions surrounding the conceptualization and design of the study and the data collectors for their diligent work in collecting and maintaining quality data (Cidália Massunda, Egness Mbanguine, Lúcia Alar, Idércia Romão, Hermínia Fernando, Nércia Vilankulo, Sara Cossa, Angela Arnaldo, Dorca Muangue). The authors also acknowledge both Inhambane Provincial and Vilankulo Rural hospitals and hospital supervisors (Raufa Mabunda, Victorino Candrinho, Cynthia Macaringue, Yolanda Messias, Fátima Nhamposse). for providing access to their patient population and hospital resources. Others in the MCMH project team also provided logistical and technical support throughout the duration of the study.

## Author Contributions

**Conceptualization**: Nazeem Muhajarine, Maud Muosieyiri

**Data Curation**: Fernanda Andre, Maud Muosieyiri, Jessie Forsyth

**Investigation**: Maud Muosieyiri, Fernanda Andre

**Statistical Analysis**: Maud Muosieyiri, Nazeem Muhajarine

**Methodology**: Maud Muosieyiri, Nazeem Muhajarine

**Supervision**: Jessie Forsyth, Ana Paula Adoni, Nazeem Muhajarine

**Fund acquisition**: Nazeem Muhajarine

**Visualization**: Maud Muosieyiri

**Writing – original draft**: Maud Muosieyiri, Fernanda Andre, Nazeem Muhajarine

**Writing – review & editing**: Maud Muosieyiri, Fernanda Andre, Jessie Forsyth, Ana Paula Adoni, Nazeem Muhajarine

## Supporting Information

**S1 Fig.** WHO MNM Abstraction Tool (WHO MNM 1.1)

**S2 Fig.** Mozambique-Canada Maternal Health (MCMH) MNM Abstraction Tool

**S3 Table.** A Breakdown of Each Sub-Category Contribution to MNM Identification Stratified by Hospital (Inhambane Provincial Hospital and Vilankulo Rural Hospital)

**S4 Table.** Results From Chi-Square Test of Independence and Kappa Statistic Between the Original WHO Disease-Criterion vs the Mozambique Expanded Disease and Comorbidities Criteria in Inhambane Provincial Hospital

**S5 Table.** Results from Chi-Square Test of Independence and Kappa Statistic Between the Original WHO Disease-Criterion vs the Mozambique Expanded Disease and Comorbidities Criteria in Vilankulo Rural Hospital

**S6 Table.** Univariable Analysis on the Association Between Maternal Characteristics and the Original WHO Disease-Criterion MNMs

**S7 Table.** Univariable Analysis on the Association Between Maternal Characteristics and the Mozambique Expanded Disease-Criterion MNMs

**S8 Table.** Univariable Analysis on the Association Between Maternal Characteristics and the Mozambique Comorbidities Criterion MNMs

**S9 Table.** Univariable Analysis on the Association Between Socio-demographic Factors and the Mozambique Comorbidities Criterion MNMs in Inhambane Provincial Hospital

**S10 Table.** Multivariable Analysis on the Association Between Socio-demographic Factors and the Mozambique Comorbidities Criterion MNMs in Inhambane Provincial Hospital

**S11 Fig.** Odd Ratios of Sociodemographic factors on MNM-defined by Mozambique Comorbidities Criterion at Inhambane Provincial Hospital

**S12 Table.** Univariable Analysis on the Association Between Maternal Characteristics and Original WHO Disease-Criterion MNMs in Vilankulo Rural Hospital

**S13 Table.** Multivariable Analysis on the Association Between Maternal Characteristics and the Original WHO Disease- Criterion MNMs in Vilankulo Rural Hospital

**S14 Fig.** Odd Ratios of Sociodemographic Factors on MNMs Defined by The WHO Disease (A), Expanded Disease (B), and Comorbidities Criteria (C) in Vilankulo Rural Hospital

**S15 Table.** Univariable Analysis on the Association Between Maternal Characteristics and the Mozambique Expanded Disease Criterion MNMs in Vilankulo Rural Hospital

**S16 Table.** Multivariable Analysis on the Association Between Maternal Characteristics and the Mozambique Expanded Disease Criterion MNMs in Vilankulo Rural Hospital

**S17 Table.** Univariable Analysis on the Association Between Maternal Characteristics and the Mozambique Comorbidities Criterion MNMs in Vilankulo Rural Hospital

**S18 Table.** Multivariable Analysis on the Association Maternal Characteristics and the Mozambique Comorbidities Criterion MNMs in Vilankulo Rural Hospital

